# Assess the Benefit of Interventional Radiologist Performing an Image Guided Biopsy

**DOI:** 10.1101/2021.04.14.21255356

**Authors:** Radu Rozenberg, Niharika Shahi, Nishigandha Burute, Ahmed Kotb, Walid Shahrour, David Kisselgoff, Christian B. van der Pol, Anatoly Shuster

## Abstract

**Purpose:** To determine whether there is any benefit in diagnostic accuracy, reduction in periprocedural complications, when image-guided biopsies are performed by interventional radiologist as compared to a general radiologist or other physician without prior special interventional training.

**Patients and Methods:** This study is a retrospective chart review of all consecutive patients that underwent imaging-guided core biopsies during one year in our hospital. Information collected included: patient age, gender, coagulation status, organ biopsied, imaging modality and equipment used, number of samples obtained, post-biopsy complications, histopathological results, and previous interventional training of a physician performing the biopsy. The quantitative data from patient charts was analyzed using a statistical program, Statistical Package for Social Sciences (SPSS).

**Results:** 449 patients were included in this study: 132 were performed using Computed Tomography guidance and 317 were performed under ultrasound guidance. The success rate of core biopsies was compared between different specialists, and was measured based on periprocedural complications, necessity of medical intervention and/or hospitalization after biopsy procedure, true positive histopathology results, and need to perform repeat biopsy of the same lesion owing to inconclusive results. Overall, IR had a success rate of 88.13%, non-IR had a success rate of 61.07%, and nephrologists had a success rate of 77.65%. The post biopsy complication rates were 5.48%, 27.52%, and 17.65% for procedures performed by IR, non-IR, and nephrologists respectively.

**Conclusion:** Our study shows an overall higher success rate and improved patient outcome when image guided biopsies were performed by IR, as compared to other non-IR physicians.

## Introduction

Procedures performed by interventional radiology specialists have become increasingly important in the management of oncologic patients [1]. Image-guided percutaneous biopsy is a standard minimally invasive procedure in oncology practice and is integral to confirming the diagnosis of cancer and determining tumor histology and grading. However, in the era of personalized medicine, where advances in knowledge of specific cellular pathways and characterization of tissue at molecular and genetic levels have resulted in an increase in targeted therapies, the role of the image-guided percutaneous biopsy and its accuracy is immensely evolving [2]. Biopsy samples are required for more than just histologic diagnosis, as the biomarker status now guides standard of care therapy in a growing number of solid neoplasms. Furthermore, biopsies are no longer being performed only at the time of initial diagnosis, rather they are being obtained at multiple time points to detect disease progression or transformation, assess treatment effectiveness, reveal residual or recurrent tumor, and thus predict the prognosis and guide next-line therapy [2]. Image-guided biopsies also play an increasing role in oncologic clinical trials [2,3]. The Food and Drug Administration (FDA) mandated the majority of targeted therapies to be accompanied by a definitive diagnostic test for appropriate patient selection [5]. Direct visualisation of the lesion enabled by image guidance during biopsy permits safe passage of a needle into an organ or mass, improving efficacy and minimising trauma to surrounding structures and risk of potentially dangerous complications such as bleeding/hematoma, vascular compromise, and hollow viscus perforation. Those minimally invasive techniques are applicable to a wide range of biopsy sites and, in most organ systems, have been reported to be highly accurate with a low complication rate [6].

There are various factors affecting the success of imaging-guided biopsies including the training of the physician performing the biopsy. Imaging-guided procedures can be performed by radiologists with a variety of training, ranging from general radiologists to fellowship-trained interventional radiologists. However, it remains unclear from the existing literature whether there is any correlation between the level of training of the physician performing the biopsy and the procedure outcomes. The likelihood of a certain specialist carrying out procedures is chiefly influenced by the availability of such a specialist in every single institution.

The choice of modality for image guidance is multifactorial and a number of options are available. US is a cheap and accessible tool, which offers the benefit of a real-time imaging allowing accurate monitoring of the needle trajectory through tissues en route to the target lesion, with the advantage of avoiding ionising radiation exposure to the patient and staff during the procedure [7]. When lesions are easily seen and reachable by US, combined with suitable equipment and sufficient operator experience, this modality can provide equivalent or superior guidance to CT [7]. However, CT guidance, in turn, enables enhanced anatomical detailing and delineation of the lesion with more precise needle localisation when compared to US [7]. Complications, if any, are more readily recognised on CT scan. It finds particular utility in thoracic, retroperitoneal, and pelvic biopsies for deeply located lesions, as well as in patients with large body habitus, for whom US would not provide an optimal visualization and guidance [8].

## Material and Methods

### Patient Selection

This study is a retrospective chart review of all patients that underwent imaging-guided core biopsies during one year in our hospital. A total of 449 adult patients underwent US guided core needle biopsy (USCNB) or CT guided core needle biopsy (CTCNB) performed by IR trained radiologists or general radiologists. Random native or transplant kidney biopsies in our institution are historically performed by a nephrologist, under US guidance with a help of a sonographer. However, IR are usually involved and assist in more challenging cases, when there is poor visualization of the kidney due to complex anatomy or patients’ large body habitus or obesity. Musculoskeletal (MSK) and breast biopsies were excluded from this study. Same CT and US equipment and room set up were used for all patients. Procedures were performed randomly by one of the two vascular interventional fellowship trained radiologists (with experience of 5 and 9 years respectively) or by general radiologists, according to their availability (with experience between 5 to 27 years). The nephrologist experience performing kidney biopsy ranges between 7 to 21 years. No residents or fellows perform biopsies in our department. The coagulation status, including INR and platelet count were obtained in all patients to exclude bleeding diathesis. All patients provided written informed consent for the procedure.

### USCNB

US examinations were performed by using an iU22 (Philips Healthcare, Best, the Netherlands) machine. After preliminary US examination, the operator performed USCNB, in accordance with the operator’s discretion. A freehand technique was used throughout the procedure to achieve accurate mass targeting. A 17-gauge coaxial needle was used by both IR and non-IR. A core biopsy was routinely performed with the matching 18-gauge cutting needle (Magnum Needles, Bard) or biopsy gun (Magnum, Bard). Nephrologists used a 16-gauge biopsy gun (Magnum Needles, Bard) without a coaxial system for random renal biopsy. Post-biopsy tract plugging using gel-foam was performed exclusively by the IR in the instances when continuous bleeding was noticed through the coaxial needle.

### CTCNB

During the procedure, a routine low-dose axial scan was obtained, with 120 kVp, 30 mAs per slice, 0.5- to 1-second rotation time, slice thickness 5 × 3.5, FOV 500 (Siemens Somatom AS +128).

No special breathing instructions were given to the patients. At our institute, we use a 19-gauge coaxial needle with a matching 20-gauge biopsy gun (Magnum, Bard) for lung biopsies and 17-gauge coaxial needle with a matching 18-gauge biopsy gun (Magnum, Bard) for other biopsies. The coaxial needle system was manually advanced in all cases.

### Data Extraction

All data were retrospectively extracted from the electronic medical record. Information collected included patient age, gender, medical history, coagulation status (INR, platelet count), organ biopsied, imaging-guidance modality, physician performing the biopsy, biopsy equipment (needle size, use of introducer needle), number of samples obtained, post-biopsy complications, and histopathological results.

### Statistical analysis

The Kruskal-Wallis test was used to compare complications between individual physicians, hospitalization rates between specialties and diagnostic biopsy rates between specialties. Chi-square test was used to assess introducer needle association with complications. Independent factors associated with complications were explored using univariate and multivariate logistic regression. Needle size and number of passes were compared to complications using the Spearman rank correlation test. Statistical analysis was defined as P<0.05. All analysis was performed using R (version 3.6.3, R Foundation for Statistical Computing, Vienna, Austria) [REF].

## Results

A total of 449 patients underwent core biopsy in the year 2017 excluding MSK, thyroid and breast biopsies (Table 1). All patients were adults (Figure 1), and a variety of organs were biopsied (Figure 2). Both IR and non-IR performed lung, liver, kidney, lymph nodes, and soft tissues core biopsies. Abdominal/pelvic intraperitoneal and retroperitoneal masses and omental biopsies were performed only by IR. Although nephrologists performed most of the random renal biopsies, IR were involved in more complicated cases, such as poor kidney visualization, complex anatomy patient’s large body habitus, obesity, etc.

**Table 1.**

| **Characteristic** | **Value** |
| --- | --- |
| Biopsies per physician (mean ± SD) | 20.4 ± 32.0 |
| Patient age (years, range) | 62 (18–91) |
| Gender | 238:211 (M:F) |
| Anticoagulation | 63:343 (Yes:No) |
| Imaging modality | 132:317 (CT:US) |
| Number of passes (median, range) | 4 (1–12) |
| Introducer needle | 272:177 (Yes:No) |
| Needle size (median, range) | 18G (14G–22G) |
| Biopsies by organ |  |
| Lung | 104/449 (23%) |
| Liver | 79/449 (18%) |
| Kidney | 110/449 (24%) |
| Lymph node | 57/449 (13%) |
| Soft tissue | 95/449 (21%) |
| Abdomen/peritoneum | 6/449 (1%) |
| Complications | 68/449 (15%) |

**Figure 1:**
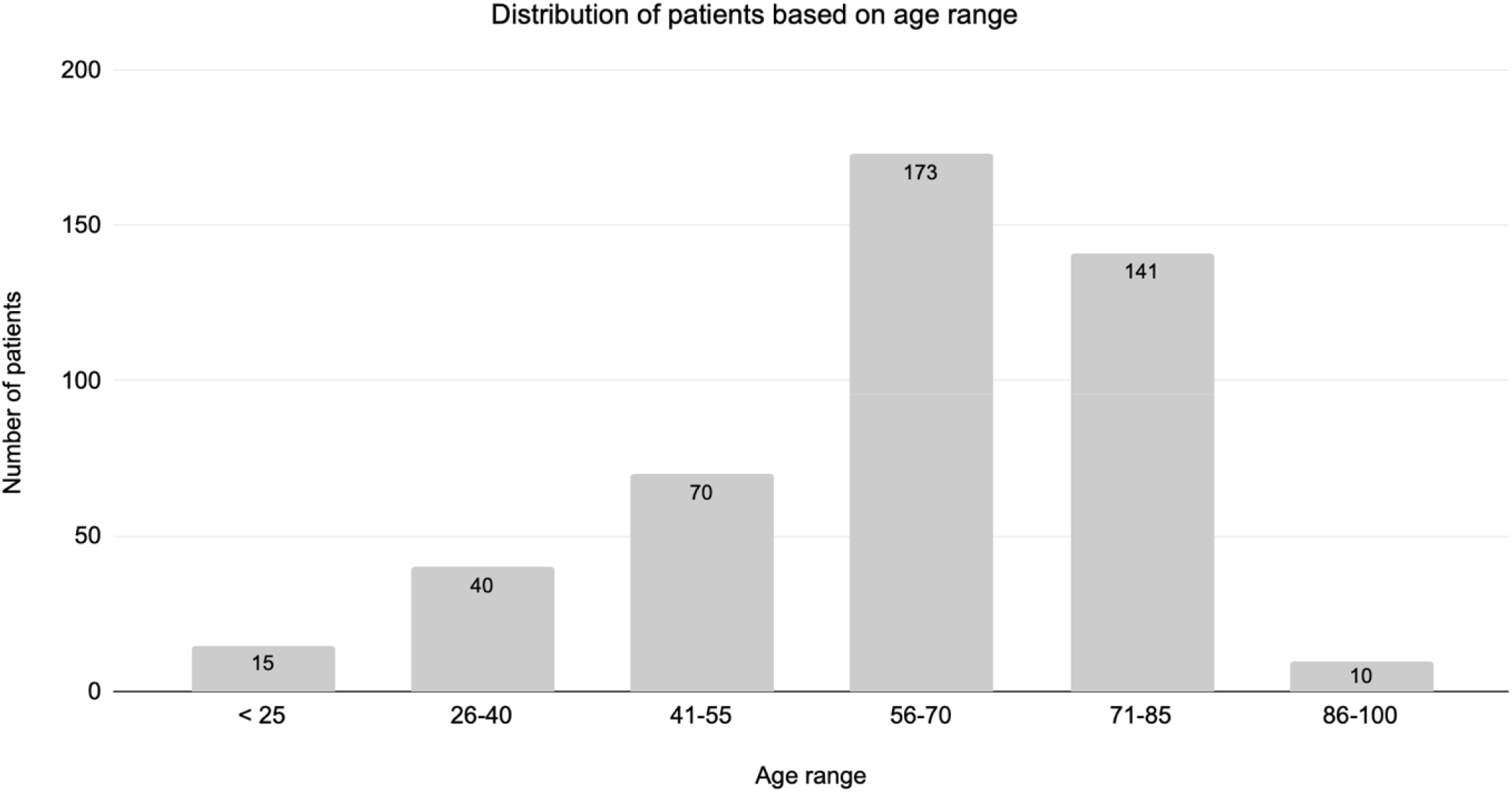
Distribution of patients based on age range

**Figure 2:**
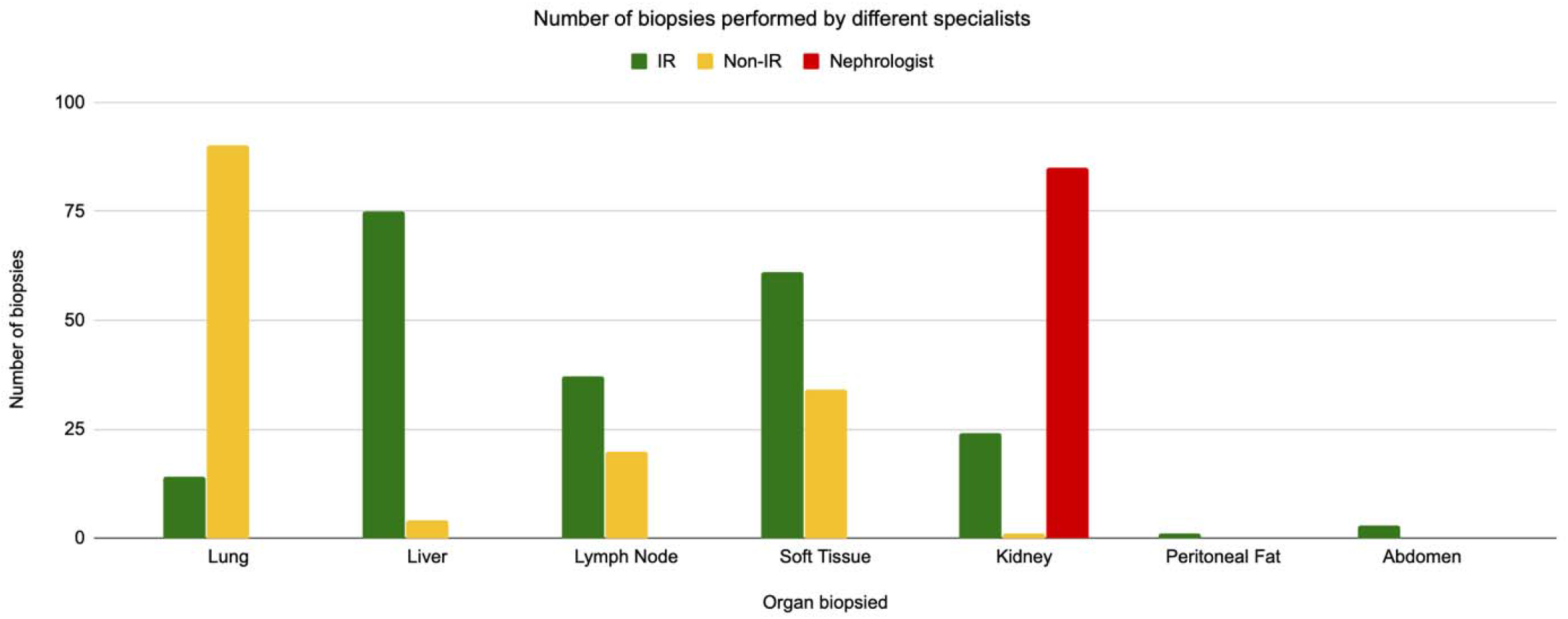
Number of biopsies performed for each organ

Complications differed significantly between physicians performing biopsies (P=0.0001). On multivariate analysis, two factors were associated with fewer complications; using ultrasound imaging-guidance as opposed to CT (P=0.0074), and soft tissue biopsy site as opposed to other organs or organ systems (P=0.0329) (Table 2). An introducer needle was used mainly by the IR and non-IR radiologists and almost never by the nephrologists (Figure 3). Post biopsy tract embolization with gel foam was used in 33 patients. In our study, none of these patients experienced any post biopsy complications. Use of an introducer needle resulted in more complications at 19% (51/272) compared to without at 10% (17/177), (P=0.0122) (Figure 3). Needle size had negligible correlation with complication rate (rho = 0.19, P<0.0001) as did the number of needle passes (rho = -0.18, P=0.0001). IR had fewer complications than the other specialties on univariate analysis (P<0.0001 and 0.0016) (Figure 4).

**Table 2.**
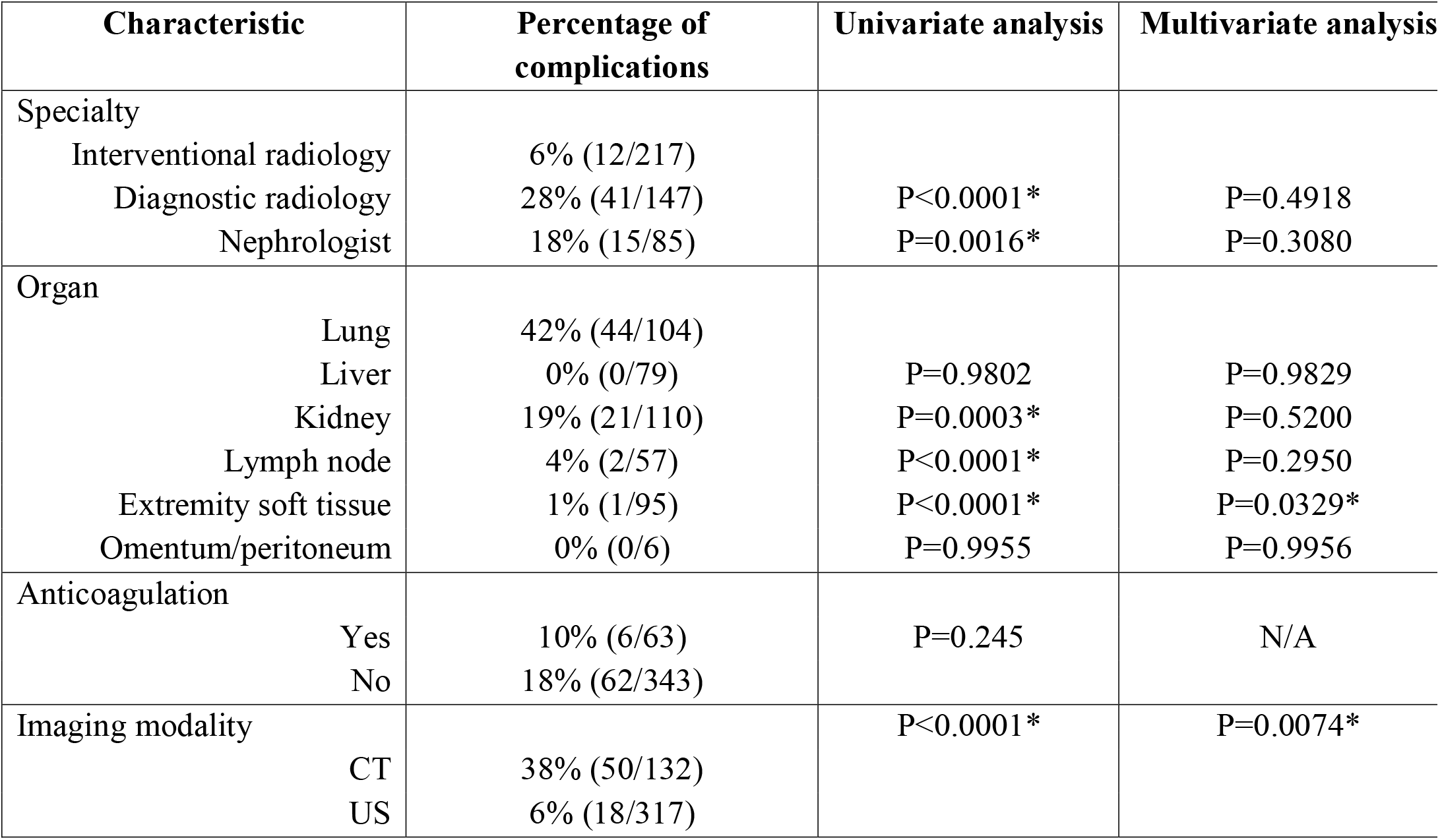

**Figure 3:**
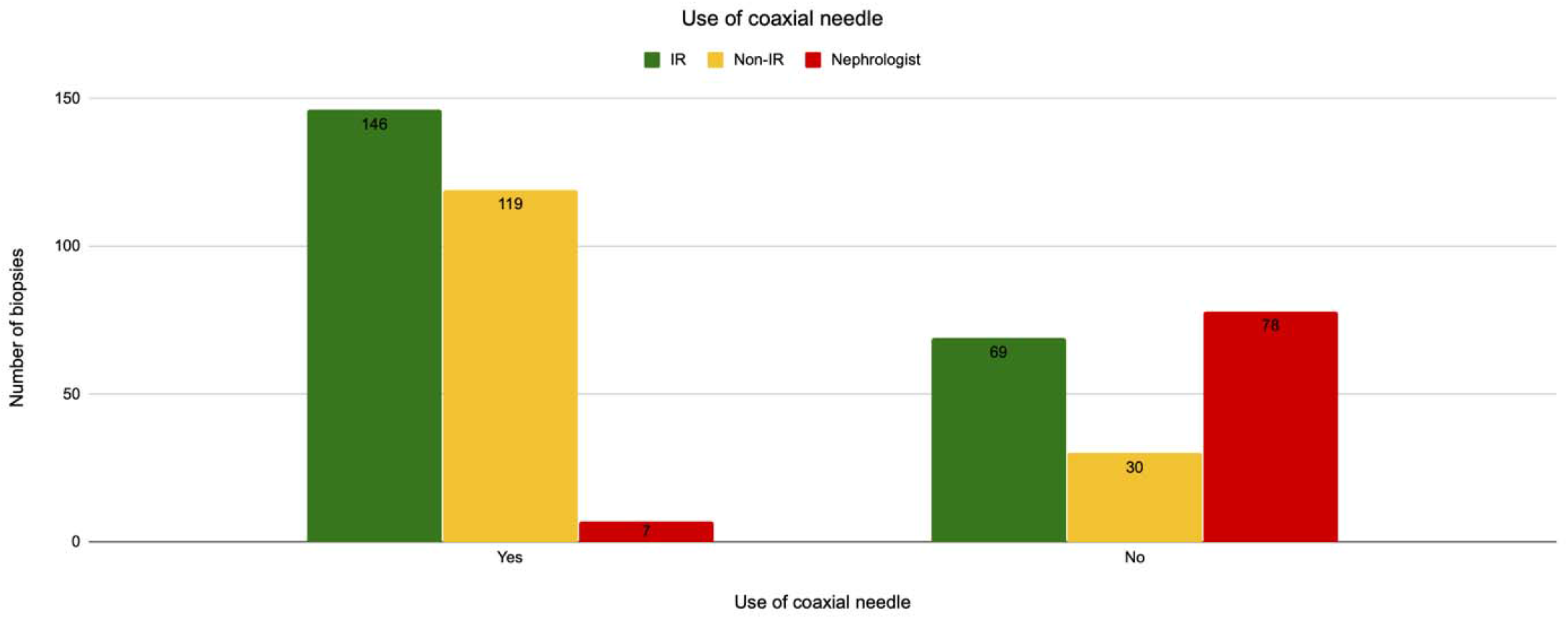
Use of introducer needle (coaxial technique)

**Figure 4:**
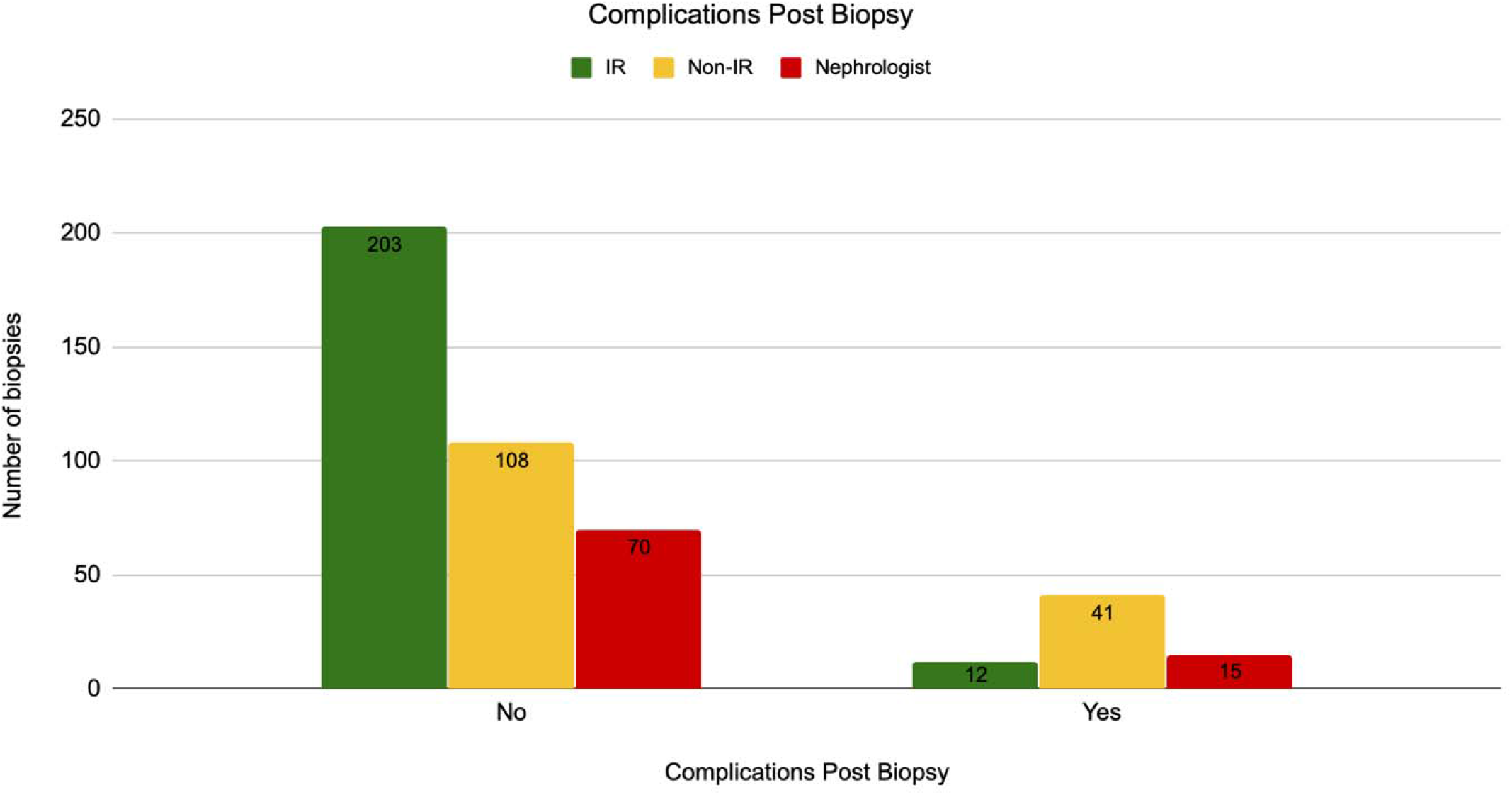
Complications post biopsy

**Figure 5:**
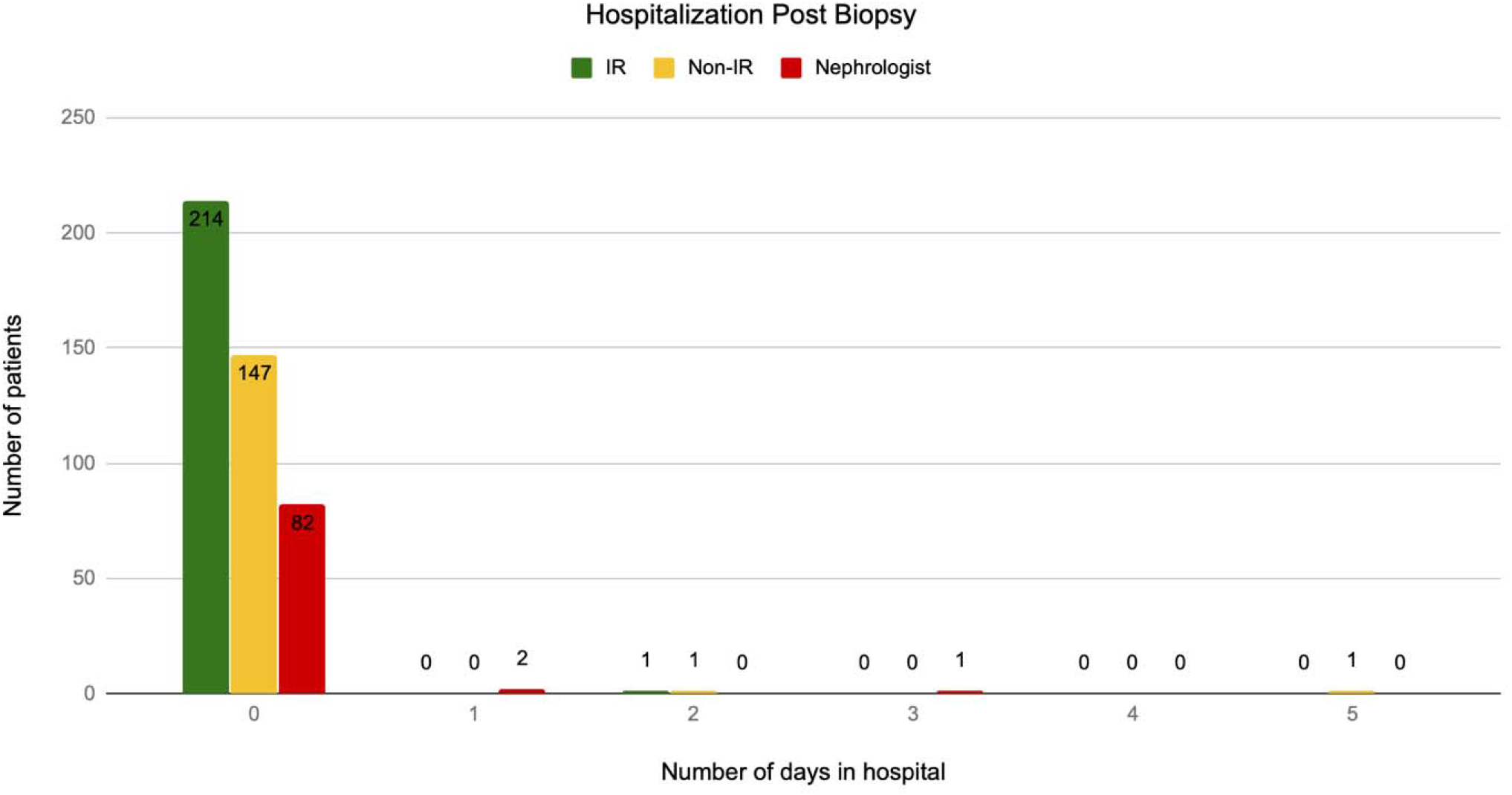
Hospitalization post biopsy

Complications from various core biopsy procedures included chest pain, dyspnea and hypoxia, hemoptysis, pneumothorax, pulmonary hemorrhage, hematuria, subcapsular and perirenal/perihepatic hematoma, intraperitoneal and retroperitoneal bleed, and soft tissue hematoma. No procedure related deaths were registered.

Most of the complications were minor and the patients did not require any interventional procedure.

Post lung biopsy, there were only 2 patients that required treatment with chest tube insertion, and they were hospitalized 2 days and 5 days respectively. Biopsy was performed on both patients by non-IR radiologist. Hospitalized patients post kidney biopsy did not require surgical or IR intervention. Hemorrhage was confirmed by CT scan and the patients were monitored in the surgical ward. Biopsy was performed by the nephrologist in all cases.

The success rate of core biopsies was compared between different specialists and was measured based on true positive histopathology results and need to re-biopsy. Overall, the IR, diagnostic radiologists, and nephrologists had success rates of 87.91%, 61.07% and 77.65% respectively which were significantly different (P=0.0001) (Figure 6). However, nephrologists performed most of the transplant kidney biopsies (73%) which may contribute to the relatively high success rate and less complications giving the more superficial location of the transplant kidney compared to the native kidney. The rest of the transplant kidney biopsies were performed by the IR, while non-IR radiologists were not involved.

**Figure 6:**
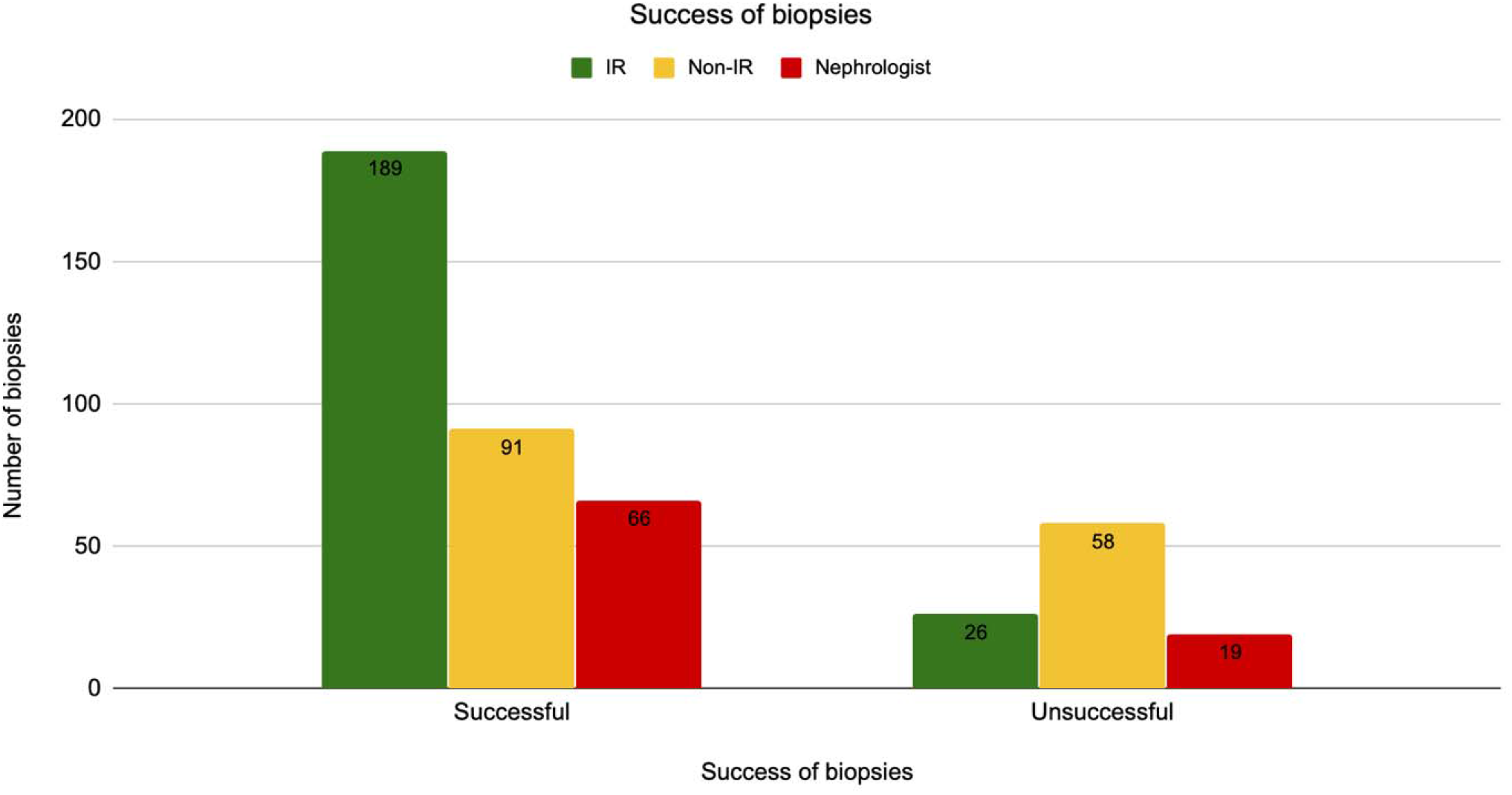
Success of biopsies

## Discussion

Image guided biopsy is routinely used for histopathological sampling of known lesions [9]. Numerous factors may affect the success of image-guided biopsies, including the training level and experience of the physician performing the procedure.

Our study illustrates an overall higher success rate when image guided biopsy was performed by fellowship trained IR, as opposed to non-IR or nephrologist, with less complications and hospitalization days. Dedicated training in performing invasive procedures, and experience in vascular and non-vascular interventions likely lead to a better choice of planning the procedure, selection of equipment and foreseeing and preventing complications. Patients undergoing imaging-guided biopsies in sites that have poor hemorrhage control are at a higher risk of complications [10, 11]. Multiple studies have shown that various periprocedural techniques, including smaller-gauge biopsy devices, use of closure tools, and post-biopsy tract embolization, all have the potential to decrease the rate of bleeding complications [10, 11, 12, 13, 14]. These techniques are routinely used by IR, but uncommonly by non-IR radiologists or other physicians, who are less familiar with the technique and equipment. Locum radiologists may also be unacquainted with the set up and tools used in a particular institution.

Limitations of this study include a retrospective chart of a single institution and only 2 IRs. Moreover, the study includes also nephrologists performing only kidney biopsies in addition to the radiologists. However, this situation reflects the reality in our institution and possibly other institutions in remote areas.

## Conclusion

Dedicated IR training may improve success rate and patient outcome in image guided biopsies compared to other non-IR physicians. Dedicated interventional training including sufficient experience in performing vascular and non-vascular minimally invasive procedures results in optimised pre-procedural planning, appropriate choice of equipment and technique, and awareness of potential complications with the expertise to prevent these in a timely manner.

In small or rural medical hospitals where IR is not available, less complicated routine procedures may be performed by other non-IR physicians. However, difficult cases with a higher anticipated rate of complications should be referred to specialized medical centers to be planned and performed by radiologists or physicians with appropriate lever of interventional training and expertise.

## Data Availability

All study data are available from the corresponding author. Data will be shared in accordance with institutional research ethics board protocols

https://docs.google.com/spreadsheets/d/1E7PlXr5Ccg0s4UEB4xWBfjtGaIFyTXvUTt3GxESp0bo/edit?ts=6025ef09#gid=1788355928

